# Predicting COVID-19 Incidences from Patients’ Viral Load using Deep-Learning

**DOI:** 10.1101/2021.08.14.21262064

**Authors:** Athar Khalil, Khalil Al Handawi, Ibrahim Chamseddine, Zeina Mohsen, Afif Abdel Nour, Rita Feghali, Michael Kokkolaras

## Abstract

The transmission of the contagious COVID-19 is known to be highly dependent on individual viral dynamics. Since the cycle threshold (Ct) is the only semi-quantitative viral measurement that could reflect infectivity, we utilized Ct values to forecast COVID-19 incidences. Our COVID-19 cohort (n=9531), retrieved from a single representative cross-sectional virology test center in Lebanon, revealed that low daily mean Ct values are followed by an increase in the number of national positive COVID-19 cases. A subset of the data was used to develop a deep neural network model, tune its hyperparameters, and optimize the weights for minimal mean square error of prediction. The final model’s accuracy is reported by comparing its predictions with an unseen dataset. Our model was the first to capture the interaction of the previously reported Ct values with the upcoming number of COVID-19 cases and any temporal effects that arise from population dynamics. Our model was deployed as a publicly available and easy-to-use estimator to facilitate prospective validation. Our model has potential application in predicting COVID-19 incidences in other countries and in assessing post-vaccination policies. Aside from emphasizing patient responsibility in adopting early testing practices, this study proposed and validated viral load measurement as a rigid input that can enhance outcomes and precision of viral disease predicting models.

## Introduction

Coronavirus disease (COVID-19) was declared as pandemic by the World Health Organization in March 2020 after the rapid global spread of the underlying severe acute respiratory syndrome coronavirus-2 (SARS-CoV-2)[1], [2]. SARS-CoV-2 can be transmitted by direct contact; within a distance of one meter through coughing, talking, or sneezing; or indirectly via infectious secretions from infected patients[3]. Therefore, the transmission rate depends on the patient’s contagious stage, viral load, and the time of exposure between individuals[4].

Reverse-transcription quantitative polymerase chain reaction (RT-qPCR) remains the gold standard for COVID-19 diagnosis[5]. RT-qPCR is a semi-quantitative diagnostic method that measures the first PCR cycle, denoted as the Cycle threshold (Ct), at which a detectable signal of the targeted DNA appears[6]. Ct value is inversely proportional to the viral load; a 3-point increase in Ct value equals a 10-fold decrease in the quantity of the viral genetic material[7]. Ct values were proposed to have potential prognostic value in predicting severity, infectiousness, and mortality among patients[8]. Ct values were also used to determine the duration an infected patient needs to quarantine[9], [10]. A high Ct value (indicating a low viral load) is detected at early stages of the infection before the person becomes contagious and at late stages when the risk of transmission is low[11]. The lowest Ct value is usually reported within three days of the onset of symptoms and co-occurs with peak detection of cultivable virus and infectivity that implies an increase in transmissibility by up to 8-folds[4]. Thus, early testing is highly recommended to promote rapid isolation practices, which can effectively interrupt SARS-CoV-2 transmission[12].

Predictive modeling of COVID-19 can determine if a rise in the number of cases is likely to occur, providing health institutions and policymakers with the opportunity to alter the trajectory of disease by preventative measures and regulations[13]. Ct is a universal parameter that is not tied to a particular demographic, and its value is correlated with transmissibility. We formalized this premise into a novel robust framework that utilizes viral load measurements, and we developed a single-feature model capable of accurately forecasting the number of new COVID-19 cases for an upcoming 7-day timeframe. The model was trained and validated on a dataset from a single cross-sectional virologic test center in Lebanon. Such a predictive model can improve the containment of the current COVID-19 pandemic and the preparedness to the threatening newly arisen SARS-CoV-2 variants that could lead for a new surge of COVID-19. Moreover, this model can assist policy makers during the vaccination stages to raise warnings about the possible adverse impact of releasing the pandemic-related restrictions. Furthermore, this model could be adapted for managing other epidemics where Ct values play a crucial role as a diagnostic indicator reflecting viral loads.

## Methods

### Study Design

This study built a predictive model for COVID-19 incidence based on Ct values using deep learning. We retrospectively collected de-identified data for all COVID-19 patients diagnosed at Rafik Hariri University Hospital (RHUH) in Lebanon between March 1, 2020, and November 31, 2020. RHUH is the country’s leading institution for COVID-19 testing and treatment, and our cohort is the largest and most comprehensive cohort in the nation[14]. Ct values were retrieved from the electronic medical database of the hospital, considering only the first positive qPCR test for each patient. The daily numbers of COVID-19 positive cases in Lebanon were obtained from the Lebanese Ministry of Public Health[15]. This study was approved by the Ethical Committee of RHUH. Written informed consent was waived since the study is retrospective and the patients’ information was de-identified.

We split the data into two groups; we used the first group (Group 1) to build the model while reserving the second group (Group 2) for validation. This approach complies with the Transparent Reporting of a Multivariable Prediction Model for Individual Prognosis or Diagnosis (TRIPOD)[16]. Inspired by clinical trial phases, the TRIPOD statement represents a classification criterion for predictive modeling. It has four types of increasing reliability. Since we split the data randomly into discovery and validation sets at the beginning of the study, our model is a TRIPOD type 2b.

### Predictive Modeling

Patients may transmit the disease before diagnosis due to interaction with other people or after it due to improper quarantining. Therefore, the future number of cases depends not only on daily Ct but also on a certain number of days in the past. The period over which past Ct values were aggregated into an input sequence for our model is defined as the sliding window *T*_1_. The forecast window *T*_2_ was fixed to 7 days throughout the study in this paper. To account for the delays between a decrease in Ct value and a rise in the number of cases, we used a recurrent neural network (RNN) model (Figure 1). A long short-term memory (LSTM) cell was used as the core building block of our RNN because of its ability to capture long-term temporal effects and trends encoded by a long sequence of inputs[17]. The LSTM network has a cell for storing temporal data and gates to control data flow and capture long-term dependencies. Each gate is composed of a multilayer perceptron[18] with *n*_hidden_ neurons that learn temporal features using backpropagation with time. We stacked 2 LSTM cells in our RNN to learn high-level feature representations (the interaction of Ct values with the past number of cases) and used a dropout probability *p*_dropout_ on the last layer to generalize better and avoid overfitting. We did not use teacher forcing during training in order to allow our model to learn from its own predictions and obtain better generalizability at the expense of training time[19]. We used an encoder/decoder paradigm to translate a variable-length input sequence (given by our sliding window) into a 7-day output sequence. All past information from the input sequence was encoded by context vectors, which were subsequently decoded by the decoder network.

**Figure 1:**
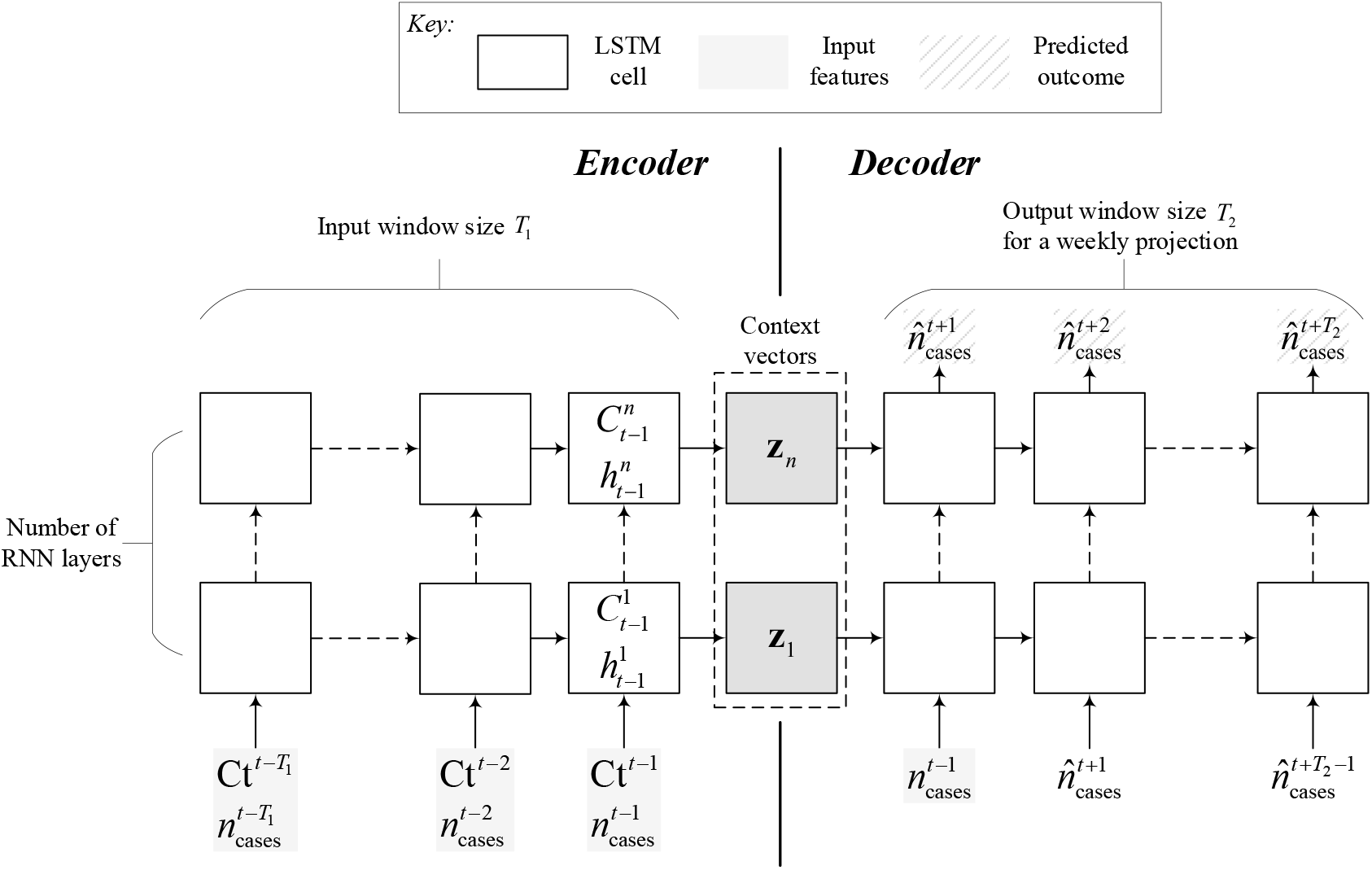
Structure of the recurrent neural network (RNN) used for forecasting the weekly number of cases. The left side of the network is the encoder that uses past information on Ct and the number of cases to create context vectors used to initialize the hidden and cell states of the decoder LSTM cells.

The network was trained using the stochastic gradient descent algorithm ADAM. We determined the optimal values of the hyperparameters (*T*_1_, *n*_hidden_, and *p*_dropout_) by minimizing the mean squared error using Group 1 only. *T*_1_ ranged from 4 to 30 days, *n*_hidden_ from 1 to 1000 neurons, and *p*_dropout_ from 0 to 1[20].

The hyperparameters and network weights were optimized using cross-validation to prevent over- and under-fitting. The cross-validation consists of outer and inner loops (Figure 2). The outer loop split Group 1 into five groups and sent four of them into the inner loop to train the model via hyperparameter optimization and internal cross-validation of the network weights. Training RNN models involves uncertainties often ignored in the literature of applied machine learning but could result in model bias. These uncertainties arise from random initialization of the weight matrices, bias vectors, dropout, and descent step sizes during backpropagation. We are the first to use a stochastic derivative-free optimization algorithm to solve such a problem via the mesh adaptive direct search, known as StoMADS[21].

**Figure 2:**
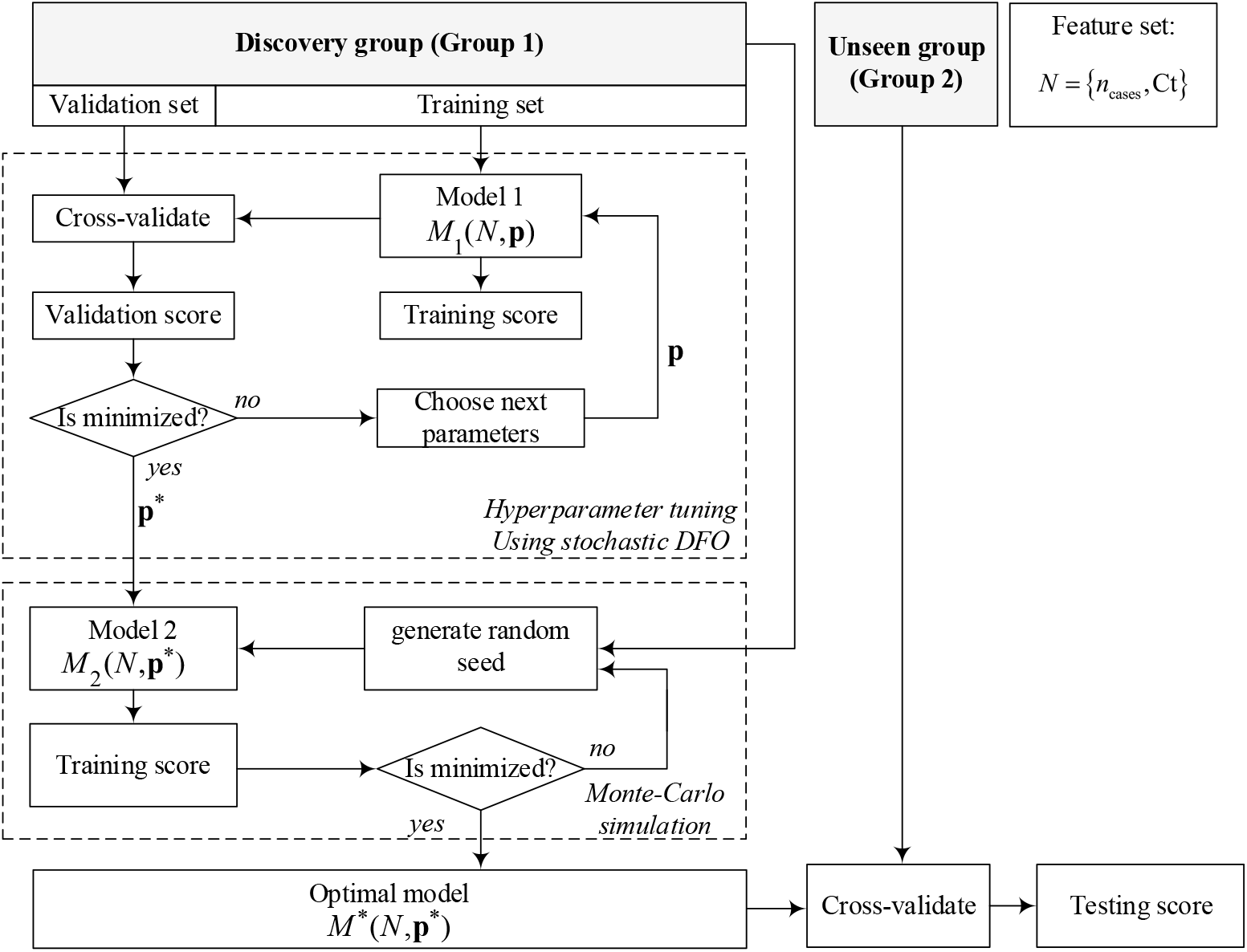
Cross-validation and hyperparameter determination scheme used for model development. Following the discovery group (Group 1), the inner loop tuned the model’s hyperparameters by minimizing the validation score using a stochastic direct search algorithm. The second loop (following tuning) generates several models randomly and bins them by training score. The best model with the lowest training score is tested on the unseen group (Group 2) to obtain the testing score.

After obtaining the optimal model in the internal loop, we scored it using the outer loop data. We then used Monte Carlo simulation to find the best possible model by randomly drawing 100 models with fixed optimal hyperparameters. The top-performing model was stored and used to make predictions for the unseen data (Group 2). The model is available on https://covid-forecaster-lebanon.herokuapp.com.

We also compared our RNN model with linear regression and support vector machine regression models that output a single value corresponding to the 7-day forward average number of cases. The linear regression model tests if the predicted 7-day average number of cases is a linear combination of mean Ct and previous number of cases, while the support vector machine regression accounts for the interaction of variables.

## Results

### Patient population

The dataset included 9,531 patients with a median age of 35 years. Figure 3 shows the bi-weekly average Ct values observed and the corresponding number of cases in Lebanon nationwide. We aggregated the individual Ct into a sequence of daily mean Ct values. Group 1 contained 5584 patients from March 6, 2020 to October 8, 2020, and Group 2 contained 3477 patients from October 9, 2020 to November 22, 2020. Both groups have comparable median age (34.5 years and 36.0 years, respectively). Group 1 was further split into five groups during model development for cross-validation: four training and one validation.

**Figure 3:**
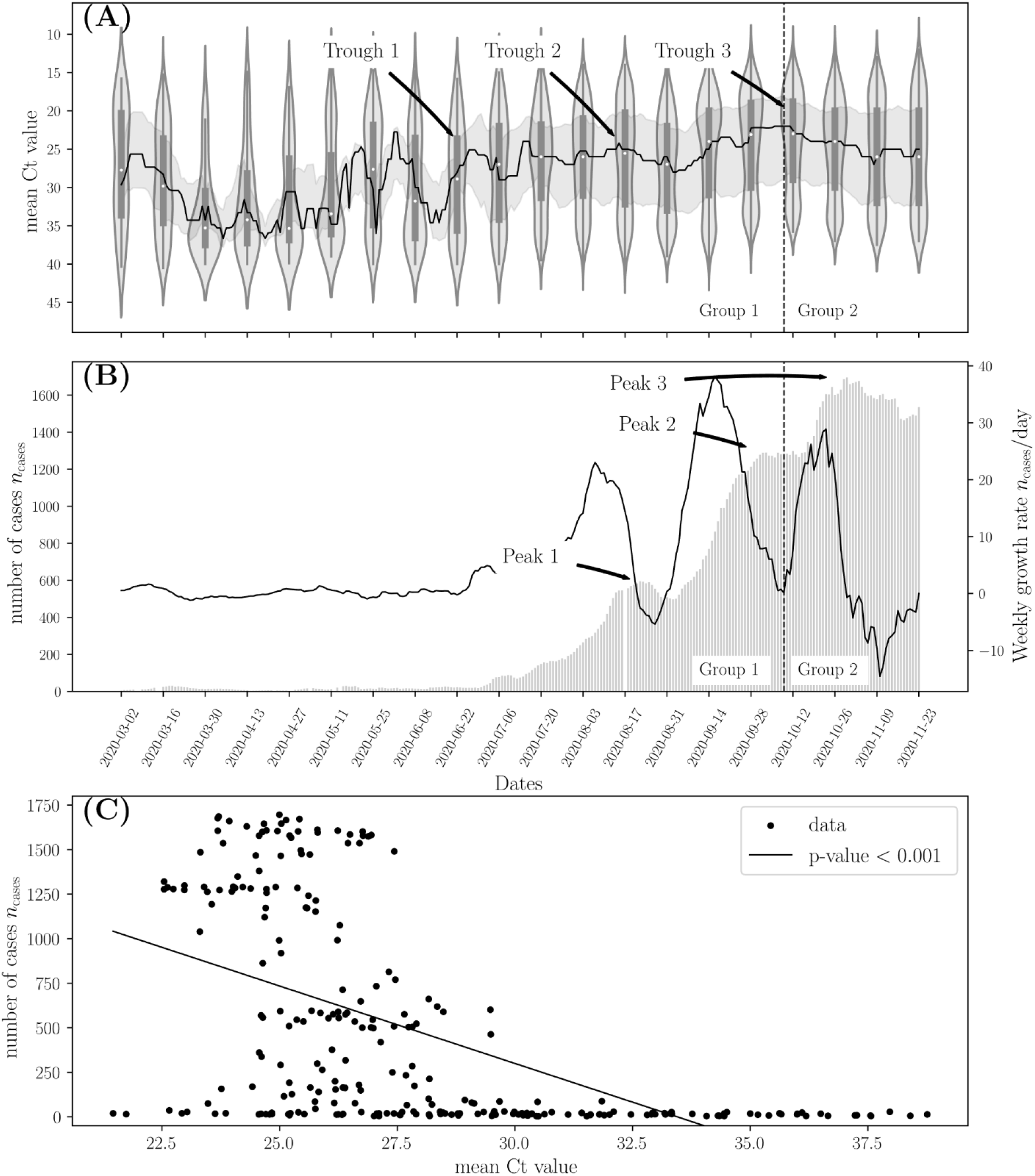
(A) Bi-weekly mean Ct values of RHUH patients. The solid line represents the median bi-weekly Ct values, and the gray shaded area represents the inter-quartile range (25-75 percentile) of the observed Ct values. (B) The grey bars show the weekly running average of the number of cases observed nationwide in Lebanon between March 1st, 2020, and December 30th, 2020 (the running average can be computed until November 23rd). The solid black line represents the growth rate in the weekly number of cases. (D) Scatter plot of biweekly mean Ct values and observed number of cases nationwide showing a clear negative value that is significant as given by p-value < 0.05

**Figure 4:**
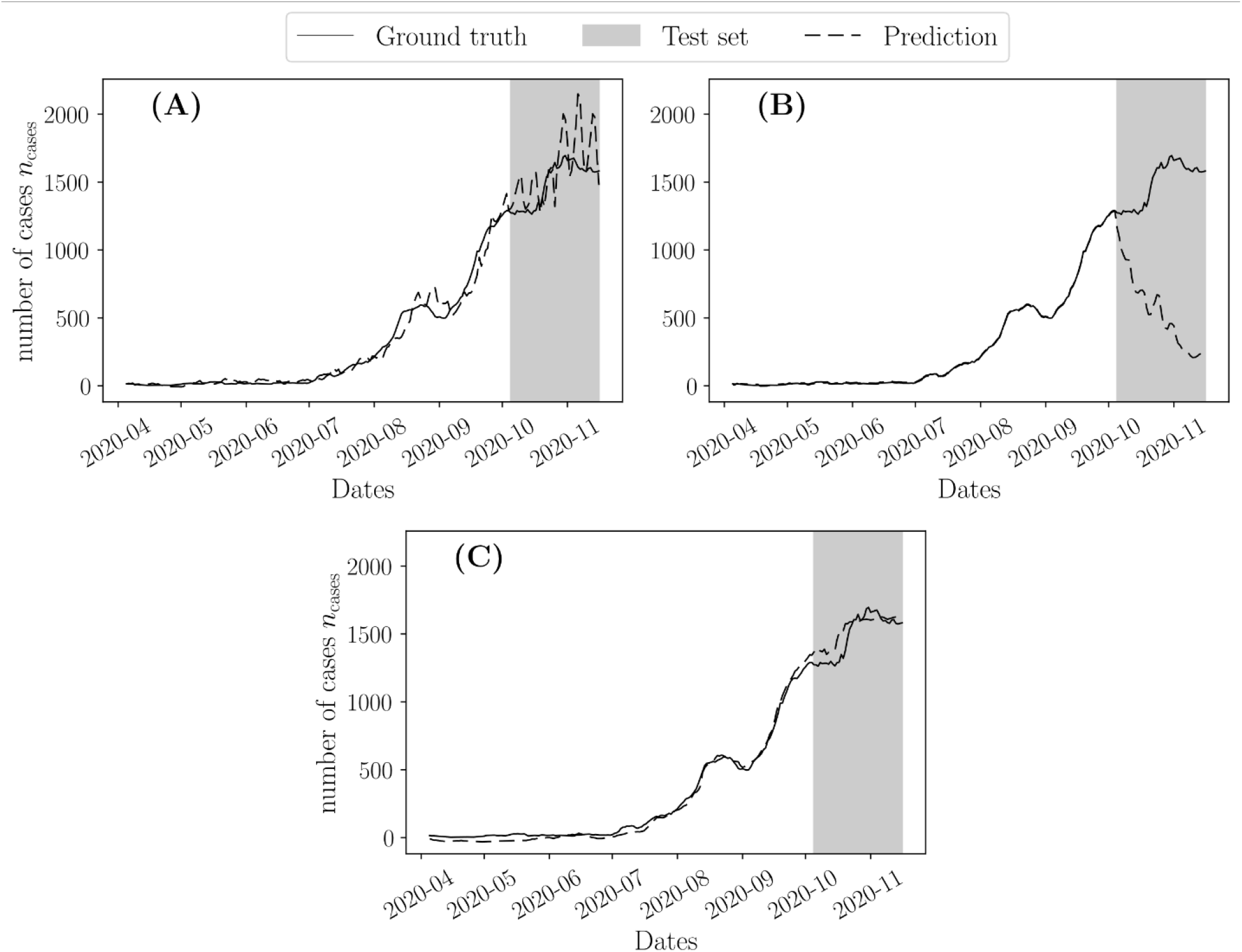
Predicted 7-day rolling average of daily number of cases using (A)a linear regression model, (B) a support vector regression model, and (C) our recurrent neural network (RNN). All models were tuned using the validation score of the discovery group. The grey shaded region represents the unseen data set (Group 2) used to test the models’ performance.

### Correlation between the national daily number of COVID-19 cases and mean Ct

We observed a lag between the incidence rate and the observed Ct values. For example, the trough in mean Ct values on October 8, 2020 (Trough 3 in Figure 3A) was followed by a peak in the growth rate in the number of cases on October 22, 2020 (Figure 3B). A peak followed this jump in growth rate in the number of cases, which plateaued on October 29, 2020, with more than 1640 cases per day (Peak 3 in Figure 3B). A similar trend was observed for peaks 1 and 2, which were superseded by troughs 1 and 2, respectively. We further observed a significant inverse correlation between mean Ct and number of cases (p<0.001), quantified by the Pearson correlation test (see Figure 3C).

### Predictive modeling

We used 3 types of predictive models to reconstruct the COVID-19 trajectory for the period of the validation set (Group 2), which was unseen during training, hyperparameter tuning, and final model selection (see Figure 5). The optimal hyperparameters for each model are listed in Table 2. The linear regression model was well calibrated on the training set (MSE ∼10^−3^) but had poor and unstable performance in the validation set (MSE ∼10^−1^). Similarly, the support vector machine regression model was well calibrated on the training dataset (MSE ∼10^−5^) but had severe underestimation of the validation set. Our RNN model showed that 29-days backward Ct values could predict the 7-day forward mean number of nationwide positive COVID-19 cases and performed well on both the discovery group (Group 1) and the unseen group (Group 2) with MSE ∼ 10^−2^ for both sets.

**Table 1:** Training and testing scores given by mean squared error of different models constructed using different feature sets. The mean squared error is computed using the standardized value of the predictions by normalizing them using the mean and standard deviation of all the daily number of cases given by 463.8 and 597.0, respectively.

| Score | Model |  |  |
| --- | --- | --- | --- |
|  | Linear regression | Support vector machine regression | Recurrent neural network (RNN) |
| Training score | $9.764 \times 10^{-3}$ | $9.385 \times 10^{-5}$ | $1.257 \times 10^{-2}$ |
| Validation score | $1.258 \times 10^{-1}$ | 3.020 | $2.316 \times 10^{-2}$ |

**Table 2:** Optimal hyperparameters of different models. The tuned hyperparameters of each model are reported on the left. The fixed hyperparameters are reported on the right

| Linear regression |  |  |  |  |  |
| --- | --- | --- | --- | --- | --- |
| Sliding window $T_1$ | | | | | |
| 4 |  |  |  |  |  |
| Support vector machine regression |  |  |  |  |  |
| Tuned |  |  | Fixed |  |  |
| Sliding window $T_1$ | | | Kernel | Regularization | epsilon |
| 15 |  |  | Radial basis function | None | 0.01 |
| Recurrent neural network (RNN) |  |  |  |  |  |
| Tuned |  |  | Fixed |  |  |
| Sliding window $T_1$ | Hidden neurons $n_{\text{hidden}}$ | Dropout $p_{\text{dropout}}$ | Learning rate | Number of epochs | Number of layers |
| 29 | 158 | 0.971 | $2 \times 10^{-4}$ | 450 | 2 |

### Deployment of the predictive model

We deployed our model in a user-friendly interface and made it publicly available through https://covid-forecaster-lebanon.herokuapp.com. The model allows the user to enter the number of cases and Ct values observed for 29 days backward (which represents the optimal sliding window obtained by the hyperparameter tuning) and the model outputs the predicted total number cases for the coming week. The data can be entered manually or copied from a spreadsheet.

## Discussion

Host viral load and the resultant Ct values have been widely proposed to evaluate the progression of SARS-CoV-2 infection and address patients’ contagiousness status[22]. Based on the premise that SARS-CoV-2 spread is highly dependent on the individual viral dynamics, we developed a model that predicts the national COVID-19 incidence rate based on mean Ct values retrieved from a single representative cross-sectional survey in Lebanon. Our COVID-19 cohort revealed that the evolution of the viral load mirrored the growth of positive national cases in the country. Low mean Ct values were followed by a large number of national positive COVID-19 cases and vice versa. To account for the effect of social interactions that could occur few days before and after testing, we used a sequence of daily mean Ct values in a stacked long-short term memory (LSTM) algorithm, a form of recurrent neural network (RNN). We trained the algorithm on a training dataset and independently validated on unseen data forming a TRIPOD type 2b model[16]. The training process utilized a cross-validation approach combined with a state-of-the-art stochastic direct search for hyperparameter tuning to prevent model over-fitting[23]. Our model showed that past Ct values obtained from a cross-sectional patient sample could be used to forecast the number of nationwide positive COVID-19 cases. We deployed our model in a user-friendly interface that allow users to predict the upcoming weekly total number of COVID-19 cases in a specific region based on the mean Ct measurements from a representative cohort.

Mathematical modeling has been widely used for predicting the course of COVID-19 pandemic. These predicting models were developed based on the applied intervention measurements and the population behavioral fluctuations, including social distancing and mask-wearing[24]. The COVID-19 reproduction number (R0), defined as the average number of naive individuals a patient can infect, has a mean estimate of 3.28 and could range from 1.4 to 6.49[25]. Although R0 can widely vary by country, culture, and stage of the outbreak, it has been used to justify the need for community mitigation strategies and political interventions[26]. So far, only few advanced and more recent models have evaluated the disease spread based on viral kinetics instead of utilizing only public health factors. Hay et al. used Bayesian inference to predict the growth rate in the daily number of COVID-19 cases as a function of Ct values[27]. They showed that the population-level Ct distribution is strongly correlated with R0 estimates in Massachusetts, USA. Although their presented method highlighted the importance of Ct for the prediction, it ignored temporal effects associated with epidemics. The temporal effects were considered only by cross-validating the model against deterministic compartmental epidemiological models, limiting their predictive ability. They estimated R0 and growth rate by using Ct to inform priors on key parameters such as the probability of infection. Hay. Et al used these estimates with parametric compartmental and Gaussian process models to predict the epidemic’s trajectory using several cross-sectional Ct samples. In comparison, our data-driven approach can directly infer the epidemic trajectory using past cases counts and Ct values. Another dataset from Bahrain demonstrated the effectiveness of Ct in predicting the epidemiological dynamics of COVID-19[28]. However, the study did not consider the interaction between different features (i.e., number of positive cases and Ct), nor does it consider temporal effects observed in epidemics. While previous predictive models relied on linear regression and Bayesian inference, our RNN model was the first to consider the temporal effects and the seasonality of epidemics that arise due to population-level dynamics. We compared our RNN model against other machine learning models that do not consider feature interaction (linear regression) and temporal effects (support vector machine regression). The RNN model outperformed the other learning algorithms since it had small and comparable training and validation errors, which demonstrates the importance of temporal relationship between cross-sectional Ct values and the nationwide number of cases.

Our dataset contained fluctuations that allowed us to extract the Ct temporal effect on the trajectory of the pandemic. Since the data came from a single institution, the fluctuations are likely to be signals in the data rather than noise. The significant changes in the Ct values in our cohort mirrored the well-recognized political, economic, and social turning points that happened in Lebanon during the pandemic. These incidences impacted the population behaviour towards COVID-19 in a consistent and well-defined manner, allowing us to track and correlate these changes with the variation in the mean Ct values and subsequently the disease spread. The early reported high mean Ct values in our cohort and the low number of COVID-19 cases in Lebanon between March 2020 and June 2020 co-occurred with a strictly imposed lockdown and a harsh awareness campaign executed by local media platforms[14]. In comparison, the sharp rise in COVID-19 cases and the decrease in mean Ct values upon diagnosis were detected after releasing the first national lockdown in July, which occurred with a significant shifting of local media attention towards the economic crisis peaking in the country. Yet, the highest jump in the number of national COVID-19 patients and the sharpest drop in Ct values were reported after the explosion of Beirut’s port in August 2020, which was classified among the most significant chemical explosions in history[29]. The devastating effects of the explosion amplified the country’s pre-existing social, economic, and health challenges, causing a significant increase in the COVID-19 positivity rate in September and November 2019, which had reached 13.9%[29], [30]. These events caused three significant peaks in the number of cases and three drops in mean Ct. Herein, we trained our model on two of these peaks and tested its ability to detect the third peak using the unseen data. Thus, our developed comprehensive training and validation scores reflect the model’s robustness against unexpected events.

The detected inversely proportional relationship between Ct values and number of national COVID-19 positive cases reflects population dynamics of transmission and demonstrates the temporal significance of Ct values. Our results emphasized the importance of early testing when patient’s viral load and infectivity is low to prompt isolation practises and thus, supress national spread of the virus. On the other hand, our newly established RNN model was able to predict precisely the upcoming one-week expected number of national COVID-19 cases based on one universal diagnostic measurement retrieved from a single representative COVID-19 cohort. Therefore, we demonstrated that the viral load measurement is a rigid input that can enhance the outcomes of disease forecasting models. Ultimately, our data promotes incorporating Ct values with other epidemiologic variables and patient demographics to predict new COVID-19 waves and to study epidemic behaviors. Our model could be used to obtain prior knowledge and insights on the virus spread patterns in other countries in the world, thus allowing more informed triage decisions and better allocation of medical resources during this pandemic. Additionally, our model can play a significant role during the vaccination process as it could detect potential rises in the number of cases after changing policies and restrictions. Finally, the use of this model could be extended to forecast other contagious viral diseases that are diagnosed by qPCR.

## Data Availability

Data will be available after the manuscript get accepted in a peer reviewed journal

## Ethics Statement

The studies involving human participants were reviewed and approved by Ethical Committee of RHUH. The ethics committee waived the requirement of written informed consent for participation.

## Data Availability Statement

The original contributions presented in the study are included in the article/supplementary materials, further inquiries can be directed to the corresponding author/s.

## Acknowledgments

The authors would like to thank the general manager of Rafik Hariri University Hospital, Dr. Firass Abiad, for his continuous support.

## Author Contributions

AK, and AA contributed to the conception of the work. KH and IC contributed to the acquisition, analysis, and interpretation of data for the work. AK and KH drafted the manuscript. AK, KH, IC, ZM RF, and MK critically revised the manuscript. All gave final approval and agree to be accountable for all aspects of work ensuring integrity and accuracy

